# Esketamine implementation challenges in psychiatry: A qualitative analysis of mental healthcare providers’ social media commentary

**DOI:** 10.1101/2025.06.01.25328722

**Authors:** Brian S. Barnett

## Abstract

**Background:** Though esketamine was approved by the Food and Drug Administration (FDA) for treatment-resistant depression in 2019, there is no published research on implementation challenges. Therefore, the aim of this study was to investigate online commentary about these challenges among mental healthcare providers.

**Methods:** Using the terms “esketamine” and “Spravato”, the author searched social media groups dedicated to mental healthcare providers on December 15, 2022. Posts and associated comments about using or implementing esketamine into practices were included in the dataset and thematically coded. Prevalence of themes about implementation challenges, illustrative quotes, and sentiment analyses are reported.

**Results:** 186 relevant posts and comments from March 12, 2019 to November 27, 2022 were identified. The most discussed challenges were billing/reimbursement (65.1%), billing codes (48.9%), staffing (18.3%), pharmacy/drug procurement (16.7%), space (11.8%), time (10.2%), “Buy and bill” acquisition (9.1%), and the FDA Risk Evaluation and Mitigation Strategy (REMS) program (7%). Sentiment regarding reimbursement for esketamine was mostly negative [72.3% (34/47 posts)], as was sentiment towards esketamine’s manufacturer (62.5% (5/8 posts). Most posts [86.7% (13/15 posts)] comparing esketamine to ketamine favored using ketamine.

**Conclusions:** These data suggest that under-reimbursement, billing challenges, and logistical barriers may be hamstringing implementation of esketamine into psychiatric practices.

## Introduction

On March 5, 2019 the United States Food and Drug Administration (FDA) approved Janssen Pharmaceutical’s esketamine nasal spray (trade name Spravato) for treatment-resistant depression (TRD) in adults when used in conjunction with an oral antidepressant^1^, and in August 2020 an indication for major depressive disorder with acute suicidal ideation or behavior was approved.^2^ Esketamine’s rapid antidepressant and anti-suicidal effects were welcomed by mental healthcare providers, though concerns were quickly raised about FDA Risk Evaluation and Mitigation Strategies (REMS) mandates, including two hours of monitoring post self-administration; excessive drug costs (as high as $6,785 for the first month of treatment); the need for additional space and staff for monitoring; and lack of long-term safety data for a drug that may require indefinite use by some patients.^3,4^ Despite these potential implementation challenges, many analysts predicted that esketamine could become a blockbuster drug, with one firm estimating potential annual sales of approximately $2.3 billion by 2024.^5^

However, these optimistic projections quickly came into question when the United Kingdom’s National Institute for Health and Care Excellence (NICE) chose not to recommend esketamine due to doubts about its clinical and cost effectiveness in 2020.^6^ The fact that sales figures for esketamine have not been reported in quarterly earnings reports^7^ released by Johnson & Johnson (Janssen is a subsidiary) suggests they are less than anticipated. Additionally, a large number of clinics employing off-label ketamine for depression continue to operate^8^, further suggesting limited clinical uptake of esketamine.

Surprisingly, even though esketamine has been on the market for four years, there is no published systematic research about potential implementation and operational challenges within psychiatry. Therefore, the aim of this study was to investigate social media discussions among mental healthcare providers about esketamine-related challenges.

## Methods

Using the search terms “esketamine” and “Spravato”, the author searched social media communities dedicated to mental healthcare providers on Student Doctor Network, r/Psychiatry Reddit forum, and Facebook on December 15, 2022. Posts and comments were included in the dataset if they discussed the author’s experience working with esketamine or implementing it into their practice.

Posts and comments not including content about implementation and delivery of esketamine treatment were excluded. Examples of excluded posts and comments include those reviewing clinical trial results for esketamine or its pharmacology, those from trainees stating whether residency programs they interviewed at offered esketamine training, and those discussing the regulatory status of esketamine as it moved through the FDA approval process for both of its currently approved indications. Relevant posts were coded for themes, and illustrative quotes were extracted. Manual sentiment analysis was conducted on quotes and descriptive statistics were used to quantify prevalence of themes within posts and associated comments. This study was declared exempt by the Cleveland Clinic Institutional Review Board.

## Results

### Quantitative analysis

There were 186 relevant posts by 83 providers in 105 unique message threads from March 12, 2019 to November 27, 2022. 123 (66.1%) posts were from Facebook, 50 (26.9%) from Student Doctor Network, and 13 (7.0%) from the r/Psychiatry Reddit forum. The most common post themes regarding challenges associated with esketamine were billing/reimbursement (65.1%), billing codes (48.9%) [a subset of billing/reimbursement], staffing (18.3%), pharmacy/drug procurement (16.7%), space (11.8%), time (10.2%), Buy and Bill drug acquisition (9.1%) [a subset of pharmacy/drug procurement], and the FDA Risk Evaluation and Mitigation Strategy (REMS) program (7%). For further details on post theme prevalence, see Figure 1.

**Figure 1.**
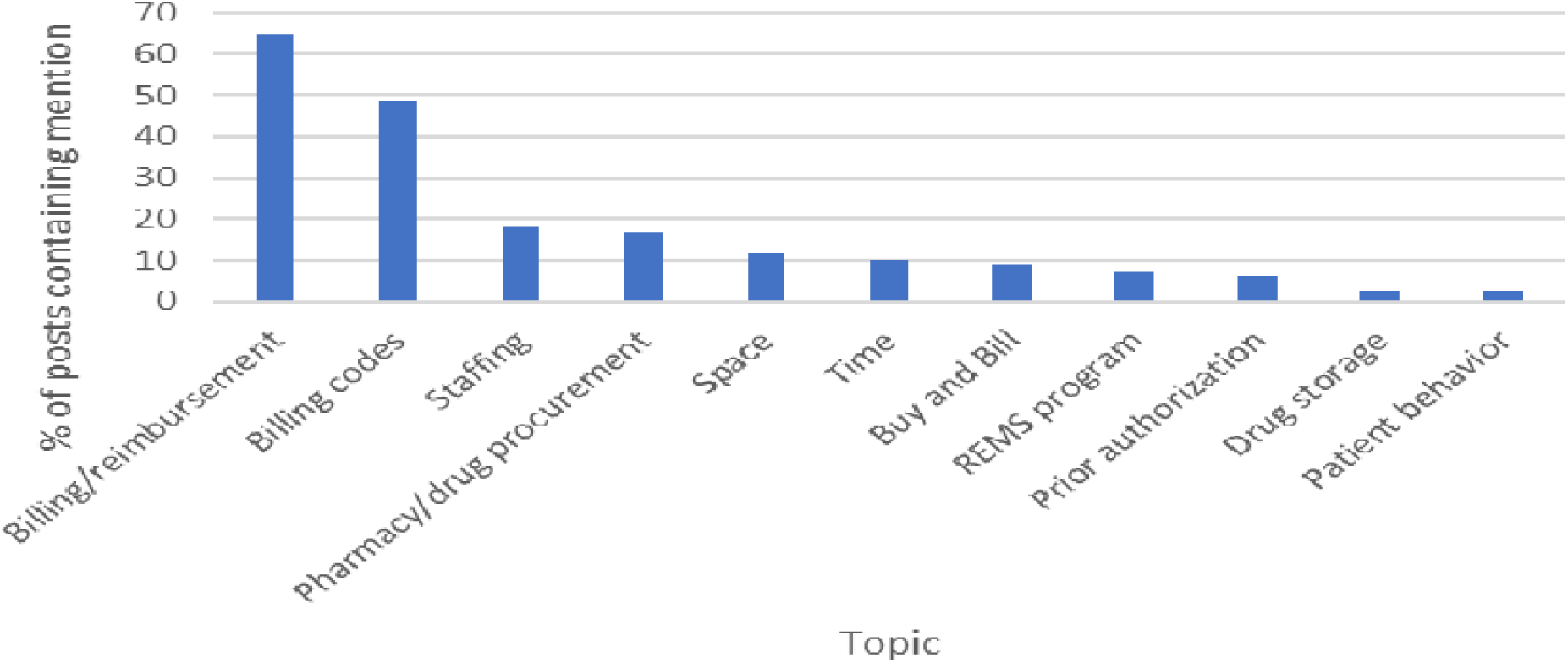
Frequency of topics mentioned in mental healthcare provider social media posts about esketamine implementation challenges

### Thematic and content analysis of posts

Here I will explore themes of discussion in social media posts discussing challenges to implementation and delivery of esketamine in psychiatric practice, while providing illustrative quotes. Particularly salient quotes for each theme are highlighted in Table 1 and not repeated in the text.

**Table 1.** Quotes emp hasizing important themes in social media posts about challenges related to esketamine implementation and delivery.

|  |  |
| --- | --- |
| **Theme** | **Select Quotes** |
| Billing code concerns |  |
|  | [Reimbursement is] a hassle and many codes are not paid for (I bill 99215 and 4 add on 15 min). Often don't get the add ons paid. |
|  | The next issue is which insurance companies will pay for the 99417? One company that is a big chunk of my panel, which I'm in the process of |
|  | dropping, doesn't cover it. Another company, also a big chunk of my panel, doesn't cover it. One tiny insurance company wants chart notes submitted with this code. |
|  | Some people bill codes for the work of the ancillary staff... I think that's getting close to jail time. |
| Staffing concerns |  |
|  | [Prior authorizations] are a huge pain and we have a full time RN doing them and coordinating with specialty pharmacies. |
|  | It was almost a full time job to manage the [esketamine patient] panel, scheduling and prior authorizations to get them started; not to mention monitoring. |
| Pharmacy and drug procurement concerns |  |
|  | Tell me how is the following scenario legal? I sent in my first script for Spravato to a REMS-certified pharmacy...insurance comes back to them as "out of network" and says the only specialty pharmacy the patient can use is Kroger Specialty Pharmacy. Well, [Kroger] isn't REMS certified. How can an insurance tell us to use a pharmacy we can't use?? |
|  | Need a reliable local pharmacy who is going to work with you on the prior authorization, delivery of medicine, etc. |
| Space concerns |  |
|  | First you need a spare room large enough for six patients, [medical assistants], and yourself. That doesn't naturally exist in the private world or someone did a bad job choosing space - increased rents. |
|  | Inpatient centers make more sense, especially in a setting of ECT [electroconvulsive therapy]. You could use mostly identical staff. Perform ECT with anesthesia and monitor Spravato patients across the room. |
| Time demands |  |
|  | Do you schedule patients every 2 hours? If yes, |
|  | means we can schedule max 4 patients for 8 hours when billing insurance? |
|  | We would have to pull providers out of their daily practice to come assess patients before and after treatment (required). And then pull them out to come assess patients PRN if something came up. |
| Buy and Bill concerns |  |
| | We have had a patient approved for esketamine, [but] the pharmacy is not REMS certified. They are asking us to buy and bill. A box is worth 10K [\$10,000]. It's really a hassle trying to get patients esketamine. |
|  | So easy to lose money with Buy and Bill Spravato. Hope is to break even and offer a service but if it bankrupts you then you can't help anyone. The cost and reimbursement have to change to be a reasonable option for all who need it. |
| REMS concerns |  |
|  | REMS is a pain to set up and you need an office administrator or nurse to fax the paperwork the day of treatment, keep log book, etc. |
| Storage concerns |  |
|  | My state is more restrictive than some but for example I can't have the medication in my clinic overnight, so if a patient does not show up the pharmacy comes and picks it up. |
| Patient behavioral concerns |  |
|  | We had [a patient attempt to drive home after esketamine], very stressful situation. The other MD kept [the patient] for 4 hours and [the patient] adamantly refused Uber/taxi. She was very alert and coherent on leaving and the MD did a lot of documentation but we also were not sure if we should call 911 to report her. |

#### Billing and reimbursement concerns

Commentary about reimbursement, including billing codes challenges, was observed in 65.1% (N=121/186) of posts and comments. Since esketamine can be billed as a medical benefit, pharmacy benefit, or either, depending on the insurer, providers expressed significant confusion about which route to use for individual insurers. One provider asked “When a patient has Cigna for their medical benefit and Caremark for their pharmacy benefit and Spravato is a drug exclusion under the pharmacy benefit, what is the next step? Must this patient be billed using Buy and Bill codes through medical benefit and will Cigna cover?”

The bulk of commentary about reimbursement focused on billing codes, which made up 48.9% (N=91/186) of all posts and comments. Billing and coding for esketamine is a complex process since different codes must be used depending on a variety of factors, including whether esketamine is a medical or pharmacy benefit for the patient, the payer, and whether the provider is purchasing esketamine directly and billing patients for it (Buy and Bill). If esketamine is supplied to the provider’s office via a pharmacy, the provider cannot report the supply of the drug on their billing claim and must use evaluation and management (E & M) codes 99202 – 99205 (new patients) and 99211 – 99215 (established patients), along with units (15 minute increments) of the 99417 prolonged office or other outpatient E & M service(s) code as necessary for reimbursement of post-administration monitoring. If the provider is supplying the esketamine directly to the patient, they can use either a J-code or S-code (non-Medicare patients only) for the esketamine in combination with the appropriate E & M codes. Further complicating matters is the fact that Medicare and some private insurers require use of a bundled G-code for esketamine and two hours post administration monitoring services instead of these J/S code and E & M code combination.

Though established by Centers for Medicare & Medicaid Services (CMS) on January 1, 2020^9^, there was little discussion in social media posts about use of the G2082 and G2083 billing codes for supervised visits for esketamine self-administration in which providers use Buy and Bill, possibly due to fewer providers using these codes because of lack of interest in participating in Buy and Bill (see below). These codes cover visits in which either esketamine doses of 56 mg or higher than 56 mg are used, respectively. Comments included:

- “G-code is okay for the BCBS [Blue Cross Blue Shield] plans except for [Medicare] Advantage. We only make $130 a treatment on those which isn’t enough to cover a room for 2 hours and staff (including me or my [physician assistant] checking in with each one).”
- “Reimbursement is actually decent with the G code.”

Since total time caring for patients receiving esketamine was reported to take between two hours and fifteen minutes and three hours, 99417 codes and difficulties in reimbursement for them were repeatedly discussed:

- One provider remarked that they use the 99215 code (40-54 minutes of care) for established patients and six units of the 99417 code but were poorly reimbursed: “We got paid [by one insurers] 16 bucks for each 99417. Janssen was expecting us to be paid $32 each 98417 but it’s falling short so far. It’s about a $200 cut from the observation codes.”
- A provider in 2021 noted, “BCBS are not covering 99417s for the rest of the year for Spravato here in [Tennessee].” Another provider wrote, “Same issue here in Alabama with our BCBS. Spravato’s only financial benefit will likely be free advertising for your clinic by [Janssen]. That and more TMS [transcranial magnetic stimulation] patients if you offer it.”
- Other providers wrote about caps on the number of 99417 codes insurers would cover, with one noting, “Some other insurances are limiting to just 2 or 3 of the extended time codes” and another adding, “I got a letter from Magellan…they seem to suggest that [reimbursement] would cap out after 3 99217 codes.”
- “Have to negotiate decent rates for those non-MD addon codes with commercial payers-that’s what we have done.”

A variety of strategies to deal with reimbursement challenges were detailed, including:

- “We charge an out-of-pocket observation fee, try to have a [physician] see the patient every time for a 99213, and often have multiple patients in the room [receiving esketamine] at once.”
- “The physician is physically present in the room sitting at a computer doing notes. The patients are all sitting in the same room, separated by individual partitions. He can view all of them throughout the 2-hour process. They have an iPad in front of them where they fill out a [Hamilton Depression Rating Scale] and [Montgomery–Åsberg Depression Rating Scale] before the treatment. Then they sit in their comfy chairs doing whatever they like for the next two hours (i.e. listen to music, relax with eyemasks, etc).”
- “I bill for time and monitor patients over video from my office down the hall. I can monitor 2 patients at a time this way and typically stagger patients 1 [hour].”
- Another provider discussed not accepting insurance for esketamine monitoring, stating, “I charge $500-$750 for 2 hours observation.”

Providers also struggled to keep up with sudden changes in payer billing code requirements for esketamine. In 2021, one provider responded to questions about whether providers were still using older billing codes, “That’s what I used last year, but apparently they are not good anymore since January 1st. Just found out. My biller is trying to find what we can use.” Another use wrote, “My billing staff informed me that BCBS will no longer accept J code for Spravato. Instead, they want S0013 to be used for Spravato administration & observation.”

Confusion over billing also produced concerns about audits by payers and legality of billing practices.

- One provider questioned the ethics of using E & M codes for esketamine monitoring: “Seems like a stretch to use time based E/M codes to justify delivering a treatment and observation after the treatment. These codes are for 1:1 evaluation or management of a physician and patient. Could probably write the note to fit the definition to justify the billing somehow but almost certainly violates the spirit/intent of those codes.”
- There was also confusion over what constituted legitimate use of extension codes, with provider stating, “I’d be worried about this too. You can bill extended time without actually seeing the patient during that time? So if I have some rating scales, [urine drug screen], and other office things, I can bill for extended time while this happens?”
- “Yeah I feel like insurance is gonna catch on to this at some point. You’re not billing some kind of group/group therapy code. You’re billing 4-6 individual notes/codes that are all occurring during the same time. Doesn’t seem like it’s gonna pass muster.”

#### Staffing concerns

Concerns about staffing were mentioned in 18.3% of posts and comments (N=34/186). Staffing issues related to prior authorization, which was mentioned in 6.5% of posts (N=12/186), were repeatedly an area of focus, with comments including:

- “Spravato requires lot of staff resources.”
- “It’s complicated to get [esketamine] approved, so you need staff for that.”
- “Getting [prior authorization] approvals can take hours of staff paid time before [treatment begins].”
- “I don’t know anyone who can afford to even have space much, less staff for this amount of money.”

Some providers reported having medical assistants [MA] monitor patients, while others had nurses do it or did it themselves. Staffing levels ranged from one MA or nurse for three to six patients. Regarding room set up and staffing, one provider stated they had, “One large room with 3 chairs separated by a curtain, and two private single rooms. MA carries a pager linked to call buttons.” Another provider remarked, “We have a nurse who handles after the MD check-in and [physician] onsite the whole time. Definitely a hassle.”

#### Pharmacy and drug procurement concerns

Pharmacy concerns appeared in 16.7% (N=31/186) of posts and comments. Providers noted challenges in working with specialty pharmacies, including delays in getting Buy and Bill applications approved by REMS certified dispensary pharmacies. The importance of a dependable pharmacy for medication delivery was repeatedly emphasized. A lack of coordination between pharmacies and insurers was recurrently discussed, with insurers sometimes requiring patients to use particular specialty pharmacies that were not REMS certified and therefore unable to supply esketamine. The merits of independent pharmacies were also highlighted. One provider remarked, “If you find the right pharmacy, they will do the PA for you. Usually an independent pharmacy, not Walgreens, etc.” Providers who worked in hospitals noted how this made esketamine treatments easier, with one commenting, “It was nice for us because we were a hospital clinic with pharmacy downstairs, so we could walk the patients there to pick [esketamine] up.”

#### Space concerns

Space concerns were mentioned in 11.8% (N=22/186) posts and comments and included:

- “[Successful esketamine programs have] big open rooms with good views, relaxing music, and staff to help [patients] do guided basic relaxation techniques.”
- “I’d consider [providing esketamine a financial] loss as you are absorbing space that could be used for more lucrative things.”
- “It’s entirely feasible to observe multiple patients at once, as others have mentioned, depending in the physical layout of your space.”
- “Psychiatry clinics typically are not equipped to do the administration and post-dose monitoring required. No prior drug has required such practices, so the infrastructure is not in place.”
- “A very large room is needed for 6 recliners spaced out plus 1 psychiatrist plus a MA running around administering doses. It might work in my waiting room, but I’d need to change the waiting room furniture out and keep the next batch of patients outside. Having a large space like this is otherwise not cost effective.”

#### Time demands

Time demands were mentioned in 10.2% of posts and comments (N=19/186). One provider wrote, “Post administration monitoring [time demands] can be a nightmare as someone has already mentioned.”

#### Buy and Bill concerns

There was also considerable confusion around the Buy and Bill program for esketamine acquisition, which was mentioned in 9.1% (N=17/186) of posts and comments. Questions included:

- “Is the intent not for the office to use buy and bill when covered under medical benefit only?”
- “What’s been your best strategy with working with REMS certified dispensary pharmacies to order for Buy & Bill? They are taking awfully long time to accept our application.”

Providers also expressed significant concern around the financial risks of Buy and Bill and reported several related mishaps. Comments included:

- “Has anyone gotten paid for buy and bill for Spravato in California? Despite giving us authorization, Magellan is now giving us a run around and not paying for treatments we have offered.”
- “That is my biggest fear about Buy and Bill, that’s why I don’t do it. If the insurance mandates this, then I don’t provide the service.”

#### REMS concerns

REMS concerns were mentioned in 7.0% of posts and comments (N=13/186). REMS requirements, including the need to monitor and document blood pressure, dissociation, sedation, and serious adverse events, were viewed as cumbersome by some providers. There was also confusion about REMS requirements, with one provider asking, “Do you know if clinics need to be registered as a pharmacy to administer Spravato that was dispensed and delivered by a pharmacy?”

#### Storage concerns

Storage concerns were mentioned in 2.2% of posts and comments (N=4/186). Comments and questions included:

- “How do you all store Spravato delivered by the pharmacy? I keep in a locked closet but wasn’t sure if we needed 2 locks or anything like that.”
- “Our clinic staff very closely documents and stores [esketamine] securely since it’s a controlled substance.”

#### Patient behavioral concerns

Patient behavioral concerns were infrequently discussed, occurring in 2.2% of posts and comments (N=4/186). These centered around patients attempting to drive home after esketamine administration, after one provider asked how they should handle this hypothetical situation. Another provider discussed how they handled this situation when it actually occurred. Two other providers reported they would terminate the patient from their practice if this situation occurred.

#### Comparisons to working with ketamine

Of 15 posts comparing working with ketamine and esketamine, 13 (86.7%) suggested ketamine was preferable to esketamine for clinical use. Provider comments in line with this point of view included:

- “Anyone get racemic ketamine compounded to be administered nasally? Cheaper than Spravato and I don’t have to follow all the regulations by REMS and fill out that annoying patient monitoring sheet at every darn visit.”
- Concerns about esketamine copays reaching as high as $250 were also discussed, including “Spravato approval is very hard and with very high copays for pts. So we mostly use generic compounded ketamine nasal spray.”
- “IV Ketamine is more efficient, cheaper and easier to control.”
- “I begin to understand why the local cowboys just order ketamine nasal sprays at compounding pharmacies instead.”
- “It’s not just the cowboys, huge organizations running residential treatment centers across the country are getting it compounded instead.”

Two comments (13.3%) noted benefit of esketamine over ketamine:

- “I think the biggest [reason esketamine is preferable] is lack of insurance coverage for ketamine for psychiatric applications…However, I have heard that some insurers have woken up and are covering IV ketamine now instead of Spravato.”
- “[With esketamine] you can get prior authorizations (private insurance no Medicare/caid) for chronic suicidality, whereas with IV ketamine we couldn’t.”

#### Reimbursement sentiment

25.3% of posts and comments (N=47/186) provided commentary on reimbursement that could be evaluated for sentiment, with 19.2% (N=9/47) being positive, 8.5% (N=3/47) neutral, and 72.3% (N=34/47) negative. Negative comments included:

- “The math still doesn’t make sense. It isn’t terrible, but it’s worse than typical billing. Spravato approval takes a lot of staff logistics with insurance. If all 6 [patients] show up at once, 6 99214’s in 2 hours is lower than I would typically bill in an insurance practice… Maybe with 6 extended time codes used multiple times simultaneously, the math is getting better, but I wonder how an audit would play out. United would see me essentially billing extended time x4 on 6 patients so 24 extended time codes in the same 2 hours. Compare that to 4 99214 + 90833 with 2 regular 99214 over 6 hours. That is a fairly typical insurance 2 hours with less logistics and probably better reimbursement.”
- “We only make $130 a treatment on those which isn’t enough to cover a room for 2 hours and staff (including me or my [physician assistant] checking in with each one).”
- “Monitoring and documenting in an office for 2 hours by a technician is a waste of space with little reimbursement.”
- “I do some esketamine in my practice, but it isn’t a lot and is more out of wanting to provide more options for patients that are treatment resistant and to do something a bit different, as it doesn’t really pay all that well for the time staff and I end up spending on it.”
- “The only $ made from esketamine is by J&J [Janssen].”
- “There is no money with Spravato, especially as an outpatient doc with low volume. We have a pretty busy Spravato program and it’s a huge money drain: requires lots of staff resources, requires space for monitoring, and you really can’t bill all that much for it.”
- “We (an academic center) offer it because our institution believes that we need to be able to offer essentially any and all treatments that are clinically appropriate, to the point that they are willing to take a financial loss to do so.”

Positive comments about reimbursement mostly revolved around relative value units (RVUs) and included:

- “The code reimbursement has been better for a while. Used to get more denials.”
- “The [successful] places I’ve seen are RVU based. One is an academic clinic in the Midwest run by an attending during their admin time. Another is a stand alone psychiatric hospital with an attached provider-based outpatient clinic. The attendings are usually inpatient doctors who see Spravato outpatients for extra RVUs… It has been incredibly profitable for the attendings and their clinic administrators do not seem to mind at all. In fact, they are using the extra Spravato RVUs as a recruiting tactic. Their typical day is inpatient rounding starting around 7am, then overseeing 4-6 Spravato patients/day while they do their notes 10:30-12:30…My hospital is building a replica model of this at our new inpatient hospital (which also has outpatient & PHP). Every attending in our group wants a piece of the pie.”
- There are several clinics in my area & also in the Midwest billing 99215 + 99417 (x4) per patient, per treatment. Comes out to 5.28 wRVU [work RVU] each treatment. Most places do 4-6 patients at a time. Physician must be physically present in the room for ∼1hr 15 min to bill for the full 2 hours. This is for private insurance and must be a “provider based outpatient clinic.” Medicare is 99215 + G2212 (x3). Comes out to about 4.6 wRVU/treatment. Swimming in wRVUs.
- “[Our reimbursement rates] did increase a lot from 2020 to 2021.”
- “I do 7 units [of 99417 extension code]. The new guidelines pay me for my check-in and my rating scales.”

#### Manufacturer sentiment

4.3% of posts and comments (N=8/186) gave opinions about Janssen Pharmaceuticals. 62.5% (N=5/8) were negative, 12.5% (N=1/8) were neutral, and 25% (N=2/8) were positive. Negative comments included:

- “And if you are using the G-code (buy and bill the drug) Janssen increased the price by about $40 a treatment, so the price of the medication further cuts the reimbursement collected.”
- “Support from Jansen not very great”
- “We just got out first “insurance investigation” back from JNJ [Janssen]. Not very helpful…just a report that basically tells us to use 99212-5 or 99202-5 for the observation, and that they need medical necessity, medical records, and [prior authorization] to get the med approved. Duh! I could have figured that out on my own.”

Positive comments included:

- “Call Spravato reps, they will come and explain the process, will help with local pharmacies, etc.”
- “[Janssen] also often did financial rebates for our patients. We worked directly with the [representatives] and they helped us organize how patients could access the treatment.”

## Discussion

This appears to be the first published systematic investigation into esketamine implementation and delivery challenges in psychiatric practice in the United States. While regulatory approval of esketamine brought hope of an effective, rapidly acting treatment for some of the 31% of patients with major depressive disorder who have TRD^10^, esketamine’s status as a schedule III substance that must be self-administered in a healthcare facility has created unique challenges to clinical dissemination. Based on this analysis, notable challenges include perceived under-reimbursement, billing and reimbursement difficulties, logistical barriers to administration, and burdens related to REMS requirements.

The most frequently discussed challenges by providers were related to billing and coding and perceived under-reimbursement by some payers for this logistically challenging treatment. Given the complexity of this treatment, the fact that esketamine can be supplied to patients either directly by providers or pharmacies, and provider hesitancy to engage in the Buy and Bill program, it is especially puzzling why Current Procedural Terminology (CPT®) codes have not been developed for esketamine monitoring services in instances where providers rely on pharmacies to supply esketamine. As a result, providers have been left to rely on the use of E and M codes, including extended time codes, which are routinely rejected by payers. Providers also seem to be experiencing significant confusion around what is required of them in terms of facetime with patients to be able to use E and M codes. In light of the recent development of Current Procedural Terminology (CPT®) codes for psychedelic drug monitoring services by psychedelic biotechnology companies in conjunction with the American Medical Association [AMA]^11^, why Janssen did not pursue or was not able to develop these codes in conjunction with the AMA is still more puzzling. The level of confusion around coding observed in this study indicates that providers would benefit from explicit guidance from payers on which billing codes to use for esketamine and what activities (including degree of facetime) are required for use of particular codes.

Most notably, this study highlights the limited ability of psychiatrists in private practice to incorporate esketamine into their practices, which is important to the field since most psychiatrists in the United States operate solo private practices.^12^ Of particular concern is the level of financial risk faced by solo psychiatrists in participating in the Buy and Bill program, in which they must pay for costly esketamine themselves and then receive reimbursement from payers, whose follow through on coverage after approving treatment appears inconsistent. While there are bundled G codes for esketamine services in cases where providers participate in Buy and Bill, these were rarely discussed in online commentary, suggesting they may primarily be used in institutional settings, where the impact of not being reimbursed for treatments after purchasing esketamine would not be as severe as for solo practitioners. Institutions may also favor the Buy and Bill program due to the potential benefits of the 340b program, which can improve profitability since it allows drug purchase discounts of 23% or more for healthcare facilities in low income areas and payer reimbursement for esketamine is based on esketamine’s average sales price.^4^

Reduction of esketamine’s pricing by Janssen could also prove beneficial for patients, providers, and the company itself by catalyzing expansion of access. From the point of approval onward concerns were raised about the price of esketamine being set too high to be a cost effective intervention, with a 2020 study concluding, “At current pricing in the U.S. market, the incremental cost-effectiveness ratio for esketamine is above the commonly cited cost-effectiveness thresholds and thus was judged to represent low long-term value for money.^13^” A recent study found that esketamine is unlikely to be cost-effective from a healthcare sector perspective, though it is less costly than ketamine from a patient perspective due to higher levels of payer coverage.^14^ Electroconvulsive therapy for patients with treatment resistant depression has also been shown to be more cost effective than esketamine.^15^ Provider discussion analyzed in this study appears to suggest that pricing of esketamine is so high as to be adversely affecting provider reimbursement to the point where providers may be resistant to using esketamine in their practice.

The FDA REMS program also appears to be the source of multiple difficulties for providers. These include the administrative burden of registering patients with the REMS system and the submission of a monitoring form at each session, as well as poor coordination between payers and REMS certified specialty pharmacies and the need to monitor patients for two hours post-administration. Multiple providers reported situations where they were required by a payer to use a particular specialty pharmacy for obtaining esketamine only to be told by that pharmacy that they did not carry esketamine because they were not REMS certified. Situations like this seem egregious and demonstrate extremely poor coordination between payers and specialty pharmacies that ultimately leaves patients unable to access a treatment that a payer claims to cover. Multiple complaints about REMS-mandated post-administration monitoring also suggest that shortening esketamine’s mandated monitoring period and allowing for clinician discretion in extending it for particular patients could make the treatment more financially viable by allowing for increased patient volume, while still preserving safety. Important adverse effects of esketamine, include dissociation, perceptual changes, and transient elevations in blood pressure, do not demand 120 minutes of post-dosing monitoring in the vast majority of patients. Dissociation and perceptual changes peak at approximately 40 minutes post-dosing and usually attenuate with repeated dosing.^16^ Metabolic variation among patients may explain why patients have varying durations of these effects.^17^ Maximum blood pressure is also typically observed approximately 40 minutes-post dosing.^16^ Importantly, recent data suggest that sublingual ketamine can safely be administered within the home via telemedicine.^18^ Furthermore, patients receiving procedural sedation with ketamine in emergency departments are routinely cleared for discharge thirty minutes post-administration due to serious adverse events being unlikely to occur after this timeframe.^19–21^ Finally, studies using intramuscular and intravenous ketamine for agitation in emergency departments and requiring vital sign monitoring for only 30 minutes post-administration have also demonstrated favorable safety findings.^22,23^ With these observations in mind, a rigid 120 minute observation period for every patient receiving esketamine, particularly during later treatments, does not appear justifiable.

Overall, data from online discussions of mental healthcare providers in this study indicate that esketamine providers feel under-reimbursed for delivering this treatment and have negative feelings towards Janssen due to costs of esketamine and perceived lack of meaningful support. Additionally, providers reported preferring to work with ketamine due to lower costs and fewer administrative burdens. This study suggests that the experience of being an esketamine provider could be improved in multiple ways by esketamine’s manufacturer, payers, and the FDA to increase access to this potentially lifesaving treatment. Future research in this area should consist of direct surveys of providers to investigate other potential challenges not captured in these data and suggested solutions for addressing implementation challenges.

### Limitations

This study is limited by the fact that only a small number of online social media forums were searched for posts and comments. Results of thematic and content analyses may have differed if comments were sourced from other forums. However, since the forums included in the search strategy are primarily used by healthcare providers, it is possible that the data in this study reflect a more realistic view of providers’ opinions due to the possibility of self-censorship in forums such as Twitter or Youtube video comments, where posts are easily accessible to the general public. Additionally, generalizability of these findings to all esketamine providers may be limited since providers who feel particularly strongly about this topic may be most inclined to discuss it online.

## Conclusions

Findings from this analysis of social media content regarding provision of esketamine treatment suggest that perceived under-reimbursement, billing and reimbursement challenges, logistical barriers, and administrative challenges associated with FDA REMS requirements may be hamstringing implementation of esketamine in psychiatric practice. Based on these observations, possible solutions to improve clinical uptake of esketamine include development of a dedicated billing code for esketamine monitoring when providers are not supplying esketamine; reduction in drug cost by esketamine’s manufacturer; streamlined coordination between specialty pharmacies and payers; and alterations to REMS requirements, including shortening of esketamine’s REMS-mandated two-hour post-administration monitoring requirement.

## Data Availability

All data produced in the present study are available upon reasonable request to the authors

